# Urinary pesticide profiles and liver disease risk in Thailand: a machine-learning risk-prediction model

**DOI:** 10.1101/2025.09.19.25336162

**Authors:** Daxesh P. Patel, Christopher A. Loffredo, Majda Haznadar, Mohammed Khan, Amelia L. Parker, Benjarath Pupacdi, Siritida Rabibhadana, Panida Navasumrit, Nirush Lertprasertsuke, Anon Chotirosniramit, Chawalit Pairojkul, Vor Luvira, Ake Pugkhem, Wattana Sukeepaisarnjaroen, Teerapat Ungtrakul, Thaniya Sricharunrat, Kannika Phornphutkul, Frank J. Gonzalez, Anuradha Budhu, Chulabhorn Mahidol, Xin W. Wang, Mathuros Ruchirawat, Curtis C. Harris, TIGER-LC Consortium

**Affiliations:** Laboratory of Human Carcinogenesis, Center for Cancer Research, National Cancer Institute, Bethesda, Maryland, USA; Georgetown University Medical Center, Washington, DC, USA; Translational Research Unit, Chulabhorn Research Institute, Bangkok, Thailand; Laboratory of Chemical Carcinogenesis, Chulabhorn Research Institute, Bangkok, Thailand; Laboratory of Environmental Toxicology, Chulabhorn Research Institute, Bangkok, Thailand; Center of Excellence on Environmental Health and Toxicology (EHT), OPS, MHESI, Thailand; Department of Pathology, Faculty of Medicine, Chiang Mai University, Chiang Mai, Thailand; Department of Surgery, Faculty of Medicine, Chiang Mai University, Chiang Mai, Thailand; Faculty of Medicine, Khon Kaen University, Khon Kaen, Thailand; Princess Srisavangavadhana Faculty of Medicine, Chulabhorn Royal Academy, Bangkok, Thailand; Chulabhorn Hospital, Chulabhorn Royal Academy, Bangkok, Thailand; Rajavej Hospital, Chiang Mai, Thailand; Cancer Innovation Laboratory, Center for Cancer Research, National Cancer Institute, National Institutes of Health, Bethesda, Maryland, USA

**Keywords:** Pesticide exposure, Hepatocellular carcinoma, Chronic liver disease, Environmental epidemiology, Risk prediction modeling, Low- and middle-income countries

## Abstract

**Background:** Building on evidence linking urinary glyphosate to chronic liver disease (CLD) and hepatocellular carcinoma (HCC), we developed urinary pesticide profiling integrated with machine learning risk prediction (MLRP) to stratify risk in high-exposure populations.

**Methods:** We conducted a case–control study within the Thailand Initiative in Genomics and Expression Research for Liver Cancer (TIGER-LC; 2011–2016; n=593): 228 CLD, 116 HCC, and 249 controls. Eight urinary pesticides were quantified by LC–MS/MS (pendimethalin, oxadiazon, metsulfuron-methyl, butachlor, 2,4-dichlorophenoxyacetic acid [2,4-D], cypermethrin, flocoumafen, bromadiolone). A composite Pesticide Load Score (PLS), with and without glyphosate, estimated burden. Two predictive models were developed: a logistic-regression Pesticide-Informed Liver Cancer Risk Score (PILCRS) and an Extreme Gradient Boosting (XGBoost) classifier that incorporated age, sex, alcohol use, occupation, and PLS. Internal validity used 1,000 bootstrap resamples with optimism-corrected calibration.

**Findings:** Predicted CLD probability increased from 30% in the lowest PLS quartile to over 70% in the highest, and HCC from 10% to 40% (p<0ꞏ0001). Relative estimates were consistent; the highest versus lowest quartile yielded odds ratios of 2ꞏ84 (95% CI 1ꞏ66–4ꞏ91) for CLD and 4ꞏ76 (2ꞏ30– 10ꞏ29) for HCC. Cypermethrin remained independently associated. After optimism correction, both models demonstrated strong discrimination and calibration.

**Interpretation:** This framework establishes a scalable, exposure-informed tool for liver disease prediction. Findings underscore pesticide burden as a modifiable risk factor and align with Sustainable Development Goal 3ꞏ9 and WHO–FAO priorities in low- and middle-income countries (LMICs). External validation is essential.

**Funding:** National Institutes of Health (USA); Thailand Science Research and Innovation.

## Introduction

Pesticide exposure is an escalating planetary health and environmental justice concern, particularly in LMICs, where regulatory infrastructure, exposure surveillance, and mitigation strategies remain critically underdeveloped. Agricultural intensification—driven by global food demand, climate adaptation, and market liberalization—has accelerated the use of hepatotoxic agrochemicals in regions where internal exposure monitoring and occupational safeguards remain limited.^1^ The implications for liver disease are substantial, given that pesticide-induced oxidative stress, mitochondrial dysfunction, and DNA damage are established pathways of hepatic injury and carcinogenesis.^2–5^

Widely used herbicides such as glyphosate, paraquat, and 2,4-D continue to be applied extensively in agriculture and home gardens despite mechanistic evidence linking them to hepatotoxicity and liver tumourigenesis.^2,4,6–8^ Glyphosate is metabolised to aminomethylphosphonic acid (AMPA) and a phosphoric acid derivative (PPA), both detectable in urine and serving as biomarkers of internal exposure, with toxicological evidence implicating them in oxidative stress, DNA damage, and hepatotoxicity.^2,9,10^ Other commonly deployed compounds—including the herbicides pendimethalin, oxadiazon, metsulfuron-methyl, butachlor, and the insecticide cypermethrin— exhibit hepatotoxic, pro-inflammatory, and fibrogenic effects in experimental models.^2,4,6,8–19^ Second-generation anticoagulant rodenticides, such as flocoumafen and bromadiolone, though less studied in humans, are environmentally persistent and induce hepatic injury via oxidative stress and coagulopathy.^6,12,19^ These compounds also contribute to groundwater contamination and biodiversity loss, compounding their ecological impact.^1,6^

Despite strong toxicological evidence of hepatotoxicity, epidemiological studies linking pesticide exposure to CLD and HCC remain scarce, often relying on indirect proxies prone to exposure misclassification and residual confounding.^2,19^ Real-world exposure typically involves chronic, low-dose contact with multiple compounds, patterns rarely captured by conventional assessment tools. The absence of internal dose surveillance and weak regulatory enforcement in LMICs further obscures population-level risk.^15,17^

Thailand exemplifies this dual burden. The country reports among the highest pesticide application rates in Southeast Asia and a rising incidence of CLD and HCC, alongside established risk factors such as chronic viral hepatitis, alcohol use, aflatoxins, and metabolic dysfunction.^1,2,12,20^ In high-intensity agricultural regions, herbicides such as glyphosate, 2,4-D, paraquat, and butachlor, and the insecticide cypermethrin, are used extensively under limited oversight.^1,2,14^ Our recent analysis from the TIGER-LC study identified significant associations between urinary exposure to glyphosate and its metabolites and increased risks of both CLD and HCC.^21^

To address these gaps, we explored a hospital-based case–control study nested within TIGER-LC. Using high-resolution LC–MS/MS, we quantified eight additional urinary pesticides and derived a composite PLS to estimate cumulative internal burden. These biospecimen-anchored metrics, combined with demographic and behavioural covariates, informed two predictive models—a logistic regression–based PILCRS and an XGBoost classifier—built and internally validated for discrimination and calibration within an MLRP framework. This interpretable, scalable framework is adaptable to artificial intelligence (AI)-enabled public-health tools for risk stratification and early prevention, aligns with WHO–FAO priorities and SDG 3ꞏ9, and underscores the need for strengthened regulation, surveillance, and policy to mitigate preventable liver-disease burden in LMICs.

## Materials and Methods

### Study Design and Participants

We conducted a secondary analysis within TIGER-LC, a multicentre, hospital-based case–control study led by the Chulabhorn Research Institute (CRI) in Bangkok and the US National Cancer Institute (NCI). Between 2011 and 2016, newly diagnosed HCC and CLD cases were recruited from five tertiary hospitals across Thailand (table 1). Hospital-based controls were recruited to approximate the age (within ±5 years), sex, and regional distribution of cases, although the groups were not fully balanced due to variations in case and control availability across sites and time. Questionnaire data and biospecimens were collected at enrollment. Detailed study design and clinical eligibility criteria have been described previously.^22,23^

**Table 1:**
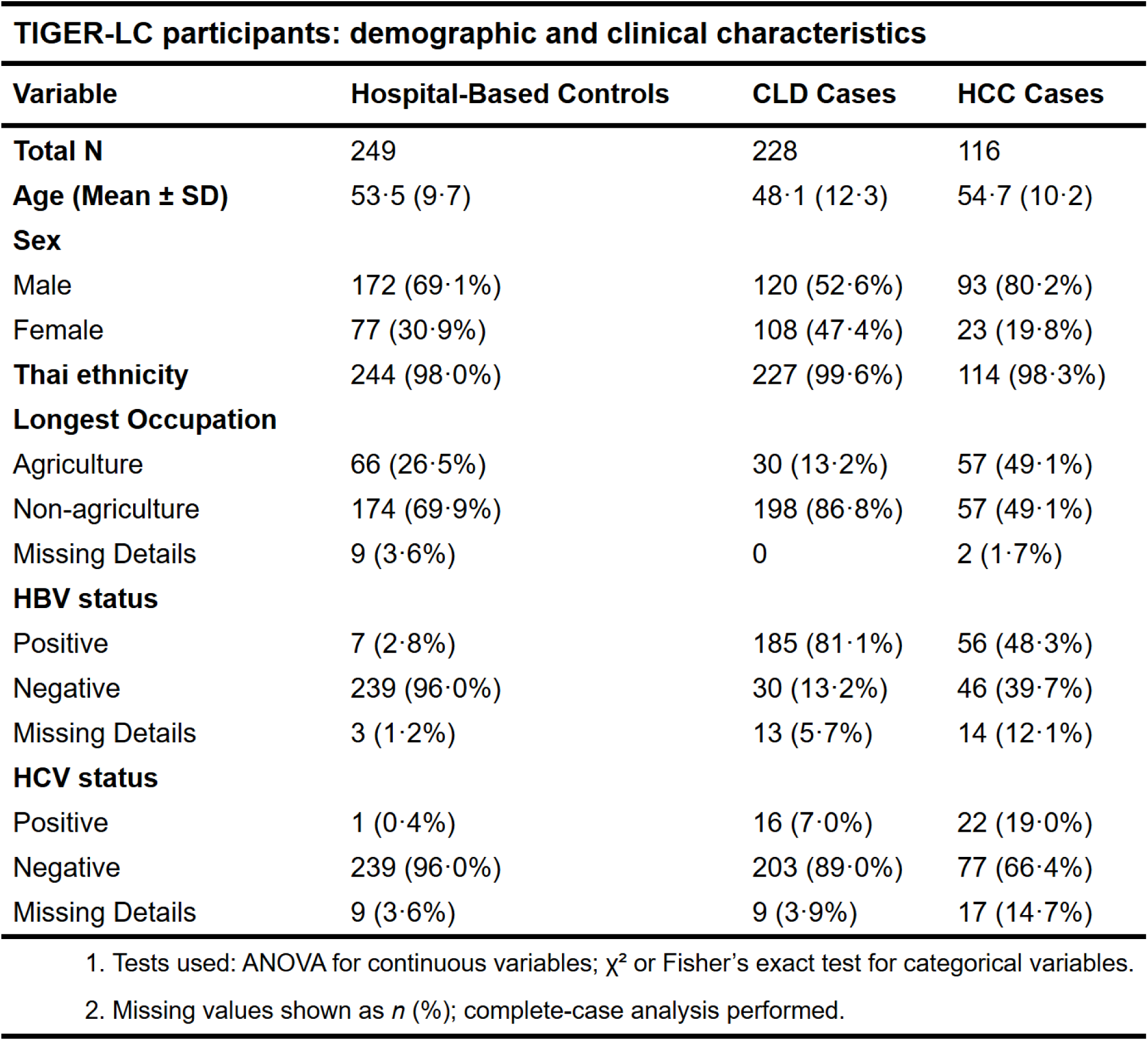
Demographic and clinical characteristics of TIGER-LC participants by disease group

### Exposure Assessment

Urinary concentrations of eight pesticides—2,4-D, pendimethalin, oxadiazon, metsulfuron-methyl, butachlor, cypermethrin, flocoumafen, and bromadiolone—were quantified using LC– MS/MS on a Waters Acquity Ultra Performance Liquid Chromatography (UPLC) system coupled to a Xevo Triple Quadrupole–Sensitive (TQ-S) micro mass spectrometer equipped with a Z-Spray™ electrospray ionization source, operated in both positive and negative ion modes. Calibration curves spanned 0.001–12.5 µM. Limits of detection (LOD) ranged from 0.16 to 467 nM, and limits of quantification (LOQ) from 0.5 to 467 nM. Matrix effects predominantly resulted in signal enhancement, and extraction recoveries exceeded 80% across analytes. Glyphosate and its primary metabolites—AMPA and PPA—were quantified separately using GC– MS, as previously described.^14^ Full assay procedures, instrument settings, and compound-specific transitions are detailed in the appendix (*Targeted LC–MS/MS assay for cross-sectional quantification of urinary pesticides: sample preparation, instrument parameters, and analytical performance*; appendix p 2) and summarised in (appendix table S1, appendix p 3).

### Expansion of Pesticide Panel and Rationale

Building on prior findings linking urinary glyphosate and its metabolites—AMPA and PPA—to increased liver disease risk.^22^ we expanded the biomarker panel to capture real-world, multicomponent pesticide exposure in Thai agricultural populations. Eight additional pesticides were selected based on participant-reported use,^9,20^ regional agricultural patterns and regulatory challenges,^9,14^ and mechanistic toxicology evidence involving hepatotoxicity, oxidative stress, and inflammatory injury.^12,20,22^ These included two herbicides, three insecticides, and three rodenticides, reflecting broad chemical class representation and plausible hepatic mechanisms (appendix table S1, appendix p 3).

### PLS Calculation

To quantify cumulative pesticide exposure burden, we constructed a composite pesticide load score (PLS) by integrating multiple urinary pesticide measurements. Two variants were defined: PLS₁₁, incorporating 11 pesticides—pendimethalin, oxadiazon, metsulfuron-methyl, butachlor, 2,4-D, cypermethrin, flocoumafen, bromadiolone, glyphosate, and its two primary metabolites (AMPA and PPA); and PLS₈, comprising the first eight of these pesticides without glyphosate or its metabolites.

All urinary pesticide concentrations were expressed in nanomolar (nM) or picomolar (pM) units and normalized to urinary creatinine, as assessed using the Jaffe method, to account for urine dilution.^14^ Calibration ranges, detection limits, and assay procedures are detailed in the appendix (*Targeted LC–MS/MS assay for cross-sectional quantification of urinary pesticides: sample preparation, instrument parameters, and analytical performance*; appendix p 2) and summarised in (appendix table S1, appendix p 3).

For each participant, the non-normalized PLS was calculated as the sum of individual analyte concentrations:

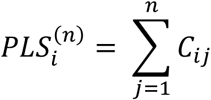

In this formulation, 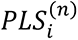 represents the cumulative PLS for participant *i*, derived from *n* compounds (n = 11 for PLS_11_ or 8 for PLS_8_). The term 𝐶_ij_ denotes the creatinine-normalised concentration of the *j*th pesticide in participant *i*, where *i* indexes study participants and *j* indexes individual analytes. This additive model captures the internal burden associated with real-world, multi-compound pesticide exposure and reflects the potential for additive or synergistic toxicological effects.^2,18,20,22,24^

To enable comparison across exposure strata, each participant’s PLS was normalized to the median score among hospital-based controls, yielding a fold-change (FC) metric:

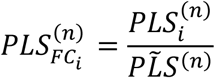

Here, 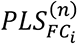 denotes the fold-change in pesticide burden for participant *i*, and 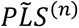 is the median PLS score across all participants or a defined reference group. These normalized scores were used to stratify exposure groups in regression analyses and machine learning–based risk prediction models.^25^

### Covariates

Covariates were selected based on established or biologically plausible associations with liver disease risk and pesticide exposure. Data were collected at enrolment using structured questionnaires and clinical assessments. Included variables were age, sex, educational attainment, geographic region, self-reported agricultural occupation, body mass index (BMI), and serological status for hepatitis B virus (HBV) and hepatitis C virus (HCV). All statistically significant covariates were included in both multivariable regression and machine learning models to account for confounding and to improve predictive performance.

### MLRP framework (PILCRS & XGBoost)

We implemented a dual-framework MLRP strategy to estimate liver disease risk associated with cumulative pesticide exposure, developing two supervised classifiers: a multivariable logistic regression model to generate the PILCRS, and a non-linear ensemble model using XGBoost.^25^ Both models included age, sex, alcohol use, self-reported agricultural occupation, and internal pesticide burden as covariates. Exposure was modelled in three forms: PLS₁₁, comprising glyphosate, its metabolites, and eight urinary pesticides; PLS₈, comprising eight pesticides only; and cypermethrin concentration as a single compound. All exposure metrics were modelled as continuous variables to preserve scale fidelity and enhance interpretability.

The binary outcome was defined as the presence of CLD or HCC versus hospital-based controls. Logistic regression models were parameterized to yield interpretable coefficients.^24^ A representative model was specified as:

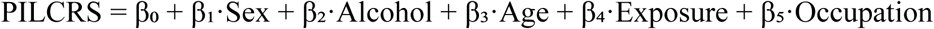

Logistic regression models were used to estimate associations between covariates and liver disease. Each β coefficient represents the change in the log-odds of liver disease per unit increase in its corresponding covariate, conditional on all other covariates in the model. Specifically, β₁ reflects the effect of sex, β₂ captures the effect of alcohol use, β₃ represents the age-associated risk, β₄ quantifies the contribution of pesticide exposure (PLS₁₁, PLS₈, or cypermethrin), and β₅ denotes the effect of agricultural occupation. The intercept (β₀) represents the model-predicted log-odds of liver disease when all covariates are held at their reference values and is not interpretable as an absolute clinical risk. In parallel, XGBoost models were optimized through grid search across key hyperparameters, including maximum tree depth, learning rate (η), subsampling ratio, and both L1 (Lasso) and L2 (Ridge) regularization; L1 penalizes the absolute magnitude of coefficients and promotes feature selection, whereas L2 penalizes the squared magnitude of coefficients and retains all features.^25,26^

### Internal Validation and Model Evaluation

Internal validation of the MLRP framework was conducted using 1,000 bootstrap resamples to assess model robustness, reproducibility, and potential overfitting.^26^ Model discrimination was evaluated using area under the receiver operating characteristic curve (AUC), and calibration was assessed using LOESS-smoothed calibration curves, bootstrapped calibration slope distributions, and the Hosmer–Lemeshow goodness-of-fit test.^26^ All models included age, sex, alcohol use, occupation, and internal exposure metrics (PLS₁₁, PLS₈, or cypermethrin) as covariates.

To evaluate predictor contributions and enhance model interpretability within the MLRP framework, Shapley Additive Explanations (SHAP) were applied to the XGBoost classifiers.^27,28^ SHAP values were used to rank variables by importance and to visualise marginal effects on predicted risk. Complete model coefficients, discrimination metrics, calibration slopes, and classification thresholds were calculated to support assessment of model performance and generalisability.

### Statistical Analysis

Analyses were conducted in R (version 4.5.0) and RStudio (version 2025.05.1). Continuous variables were summarized as mean ± standard deviation (SD); categorical variables as counts (%). Group differences were assessed via Kruskal–Wallis and chi-squared tests; pairwise comparisons used Dunn’s test with Bonferroni correction. Multivariable logistic regression and XGBoost were implemented as components of the MLRP framework to evaluate exposure– outcome associations. All p-values were two-sided; p < 0.05 denoted statistical significance. Analyses and data presentation adhered to Strengthening the Reporting of Observational Studies in Epidemiology (STROBE),^26^ and Transparent Reporting of a Multivariable Prediction Model for Individual Prognosis or Diagnosis (TRIPOD) guidelines.^27^

### Ethics Approval

The study was approved by the institutional review boards of all participating institutions. Written informed consent was obtained from all participants prior to enrollment.

## Results

### Study Population Characteristics

Among 593 participants, 228 had CLD, 116 had HCC, and 249 were hospital-based controls (table 1). Compared with hospital-based controls, CLD cases were younger and HCC cases older, with significant differences in sex, occupation, and HBV/HCV status (all *p*<0ꞏ001). Thai ethnicity was uniformly prevalent and did not differ significantly across groups (*p*=0ꞏ38).

### Pesticide Profiling and Exposure Patterns

Urinary concentrations of 11 analytes—including glyphosate, its metabolites AMPA and PPA, and eight additional pesticides—were quantified using LC–MS/MS and GC–MS, with quality control including matrix-effect testing, recovery validation, and creatinine normalisation (appendix table S1, appendix p 3). Radar plots illustrated multidimensional exposure profiles across all analytes, with consistently higher concentrations in CLD and HCC compared with hospital-based controls (figure 1A). Individual distributions of seven additional pesticides are presented in (appendix figure S1, appendix p 5).

**Figure 1:**
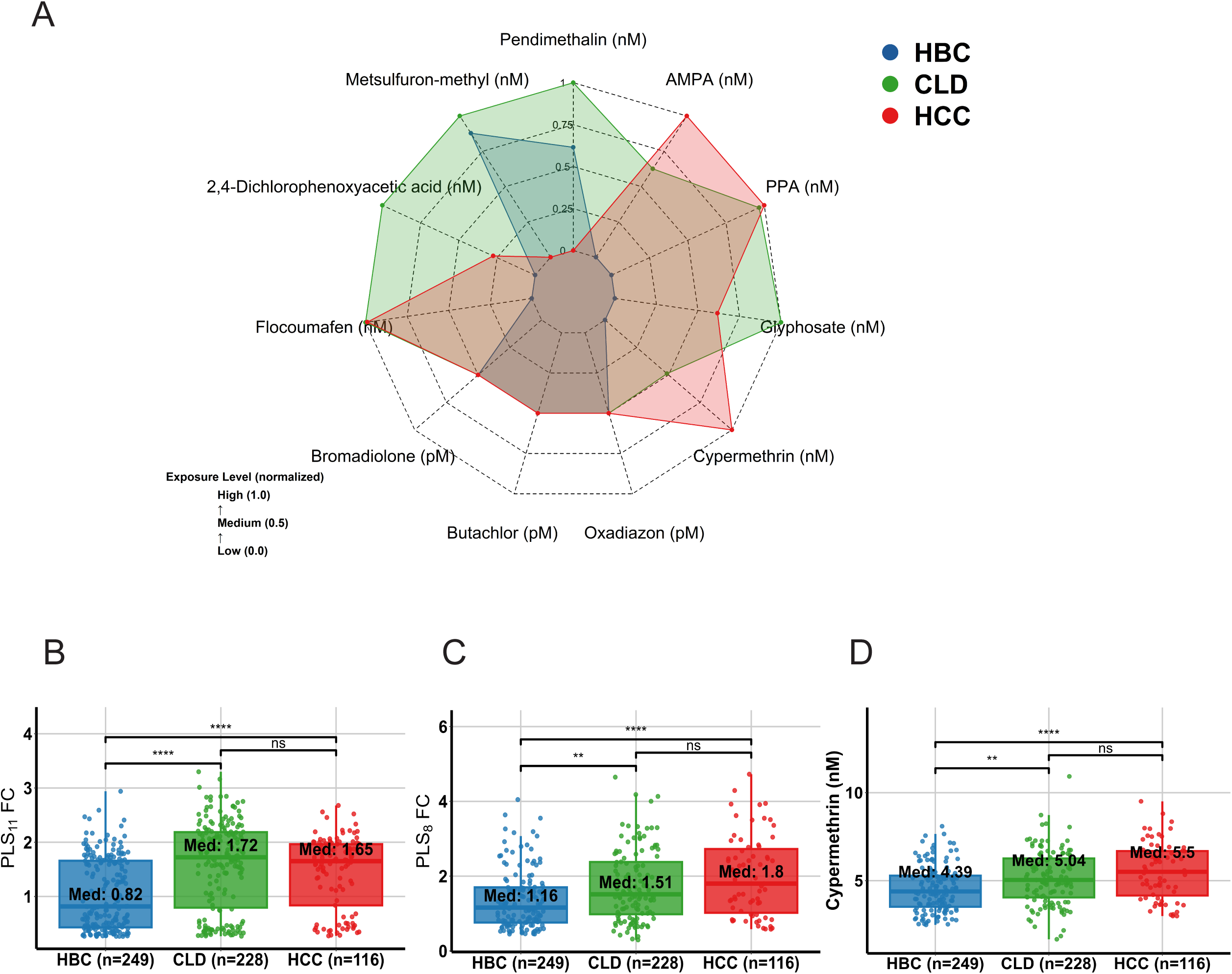
Urinary pesticide concentrations and exposure scores by disease group. (A) Radar plot shows scaled median urinary concentrations of 11 pesticides across hospital-based controls (HBC), chronic liver disease (CLD), and hepatocellular carcinoma (HCC) groups. (B–D) Boxplots display groupwise distributions of PLS₁₁ fold-change (B), PLS₈ fold-change (C), and cypermethrin (nM) (D) in HBC, CLD, and HCC groups. Asterisks denote significance by Wilcoxon rank-sum test; *ns* = not significant; Median values shown within boxes.

### Composite Exposure Metrics and Cypermethrin Stratification

Two composite indices were derived: PLS₁₁, comprising glyphosate, its metabolites, and eight urinary pesticides; and PLS₈, comprising the eight pesticides alone. Both were scaled as fold-changes relative to hospital-based control medians and were significantly elevated in CLD and HCC (figure 1B–C). Cypermethrin was analyzed separately because it dominated the exposure distribution and was markedly higher in CLD and HCC than in hospital-based controls (figure 1D).

### Multivariable Regression Analysis

Multivariable logistic regression adjusted for age, sex, alcohol use, and occupation showed that PLS₁₁ and cypermethrin were significantly associated with higher odds of disease, whereas PLS₈ showed weaker associations. For PLS₁₁, adjusted ORs for the highest quartile were 2ꞏ84 (95% CI 1ꞏ66–4ꞏ91) for CLD and 4ꞏ76 (2ꞏ30–10ꞏ29) for HCC (table 2). For PLS₈, ORs were 1ꞏ54 (0ꞏ89– 2ꞏ69) for CLD and 1ꞏ76 (0ꞏ92–3ꞏ40) for HCC (table 2). For cypermethrin, ORs were 1ꞏ88 (1ꞏ04– 3ꞏ46) for CLD and 3ꞏ26 (1ꞏ70–6ꞏ34) for HCC (table 2). Sensitivity analyses excluding HBV/HCV-positive participants and dose–response trends supported these findings. In fully adjusted models, high versus low PLS₁₁ exposure was associated with ORs of 2ꞏ01 (95% CI 1ꞏ18–3ꞏ47; *p*=0ꞏ0115) for CLD and 1ꞏ80 (1ꞏ21–2ꞏ70; *p*=0ꞏ0042) for HCC (appendix figure S2, appendix p 6). For cypermethrin, ORs were 3ꞏ53 (1ꞏ68–7ꞏ79; *p*=0ꞏ0012) for CLD and 1ꞏ69 (0ꞏ99–2ꞏ90; *p*=0ꞏ0530) for HCC. No effect modification was observed by HBV/HCV status or occupation. Among CLD cases, males had higher PLS₁₁ values than females (*p*<0ꞏ001).

**Table 2:**
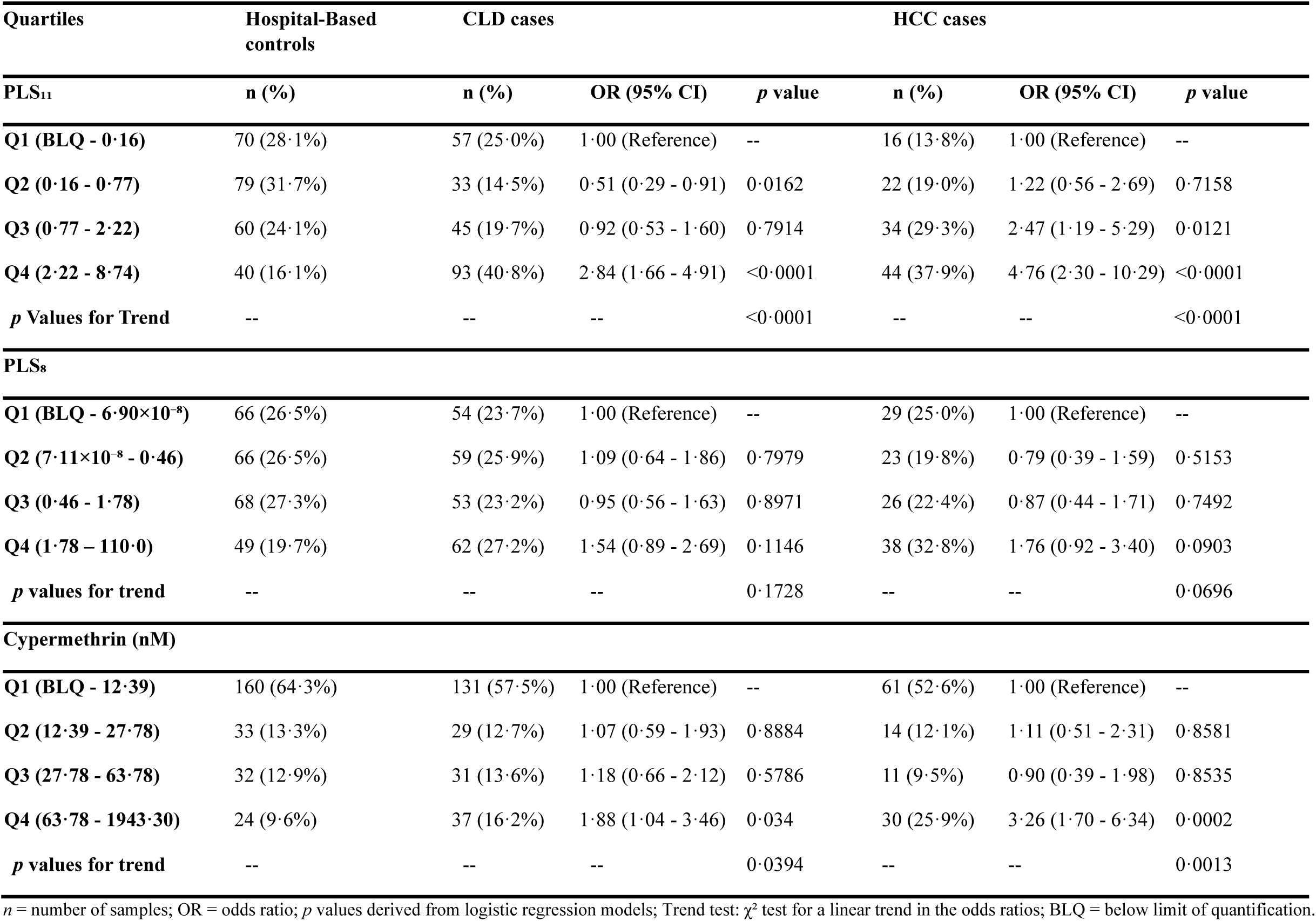
Odds ratios for chronic liver disease and hepatocellular carcinoma by quartiles of pesticide exposure metrics

### Predictive Modelling, Classifier Performance, and Predicted Risk Probability

To evaluate predictive performance within the MLRP framework, three logistic regression–based PILCRS models (PILCRS₁₁, PILCRS₈, and PILCRS_CYP_) and parallel XGBoost classifiers were developed (figure 2A–B). PILCRS₁₁ showed the highest discrimination among logistic models (AUC 0ꞏ85 for CLD, 0ꞏ89 for HCC), followed by PILCRS₈ (0ꞏ83 and 0ꞏ87) and PILCRS_CYP_ (0ꞏ78 and 0ꞏ84). XGBoost using PLS scores or cypermethrin exposure plus covariates achieved AUCs of 0ꞏ86 for CLD and 0ꞏ91 for HCC, with Brier scores of 0ꞏ09 and 0ꞏ07, indicating excellent calibration. Risk score distributions showed clear separation between cases and hospital-based controls, most pronounced for PILCRS₁₁ and PILCRS_CYP_ (figure 2A–B). Stratified curves demonstrated progressive increases in predicted risk probabilities with higher scores (figure 2C): for PILCRS₁₁, CLD rose from 45% in Q1 to 70% in Q4 and HCC from 10% to 21%; for PILCRS₈, CLD from 47% to 53% and HCC from 4% to 9%; and for PILCRS_CYP_, CLD from 25% to 59% and HCC from 5% to 15% (figure 2C).

**Figure 2:**
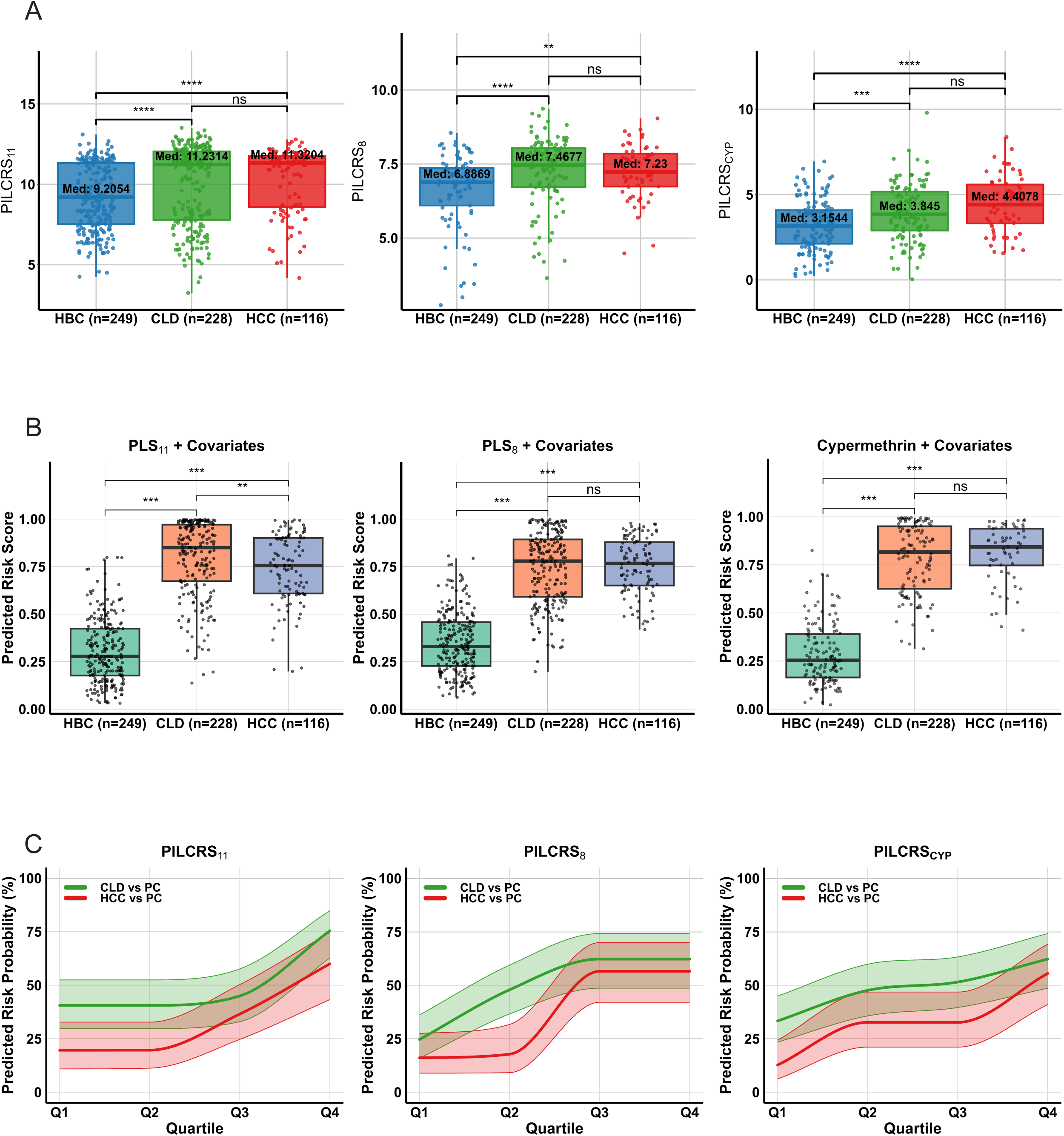
Predictive liver disease risk scores derived from PLS + covariates models by disease group. (A) Boxplots show distributions of PILCRS₁₁, PILCRS₈, and PILCRS_ᴄyp_ across HBC, CLD, and HCC, derived from logistic regression models incorporating PLS scores or cypermethrin plus covariates. (B) Predicted liver disease risk scores from logistic regression models using PLS₁₁, PLS₈, or cypermethrin plus covariates. (C) Quartile-based predicted risk probabilities for PILCRS₁₁, PILCRS₈, and PILCRS_ᴄyp_, showing monotone increasing estimates with 95% confidence intervals for CLD vs PC and HCC vs PC. Boxes indicate medians and IQRs; horizontal bars show Wilcoxon rank-sum test results; *ns* = not significant.

### Model Calibration, Interpretation, and Internal Validation

All models showed acceptable calibration, with Hosmer–Lemeshow tests non-significant (*p* > 0ꞏ10). Calibration plots from PLS plus covariates models (figure 3) showed good agreement between observed and predicted probabilities across PILCRS₁₁, PILCRS₈, and PILCRS_CYP_. Bootstrap validation (1000 resamples) confirmed robustness, with optimism-corrected AUCs of 0ꞏ83–0ꞏ90 and calibration slopes of 0ꞏ73–0ꞏ90. Distributions of slope estimates indicated minimal overfitting, and bootstrap confidence intervals supported stability (appendix figure S3, appendix p 7). Discrimination was strong, with consistent case–hospital-based control separation (appendix figure S4, appendix p 8).

**Figure 3:**
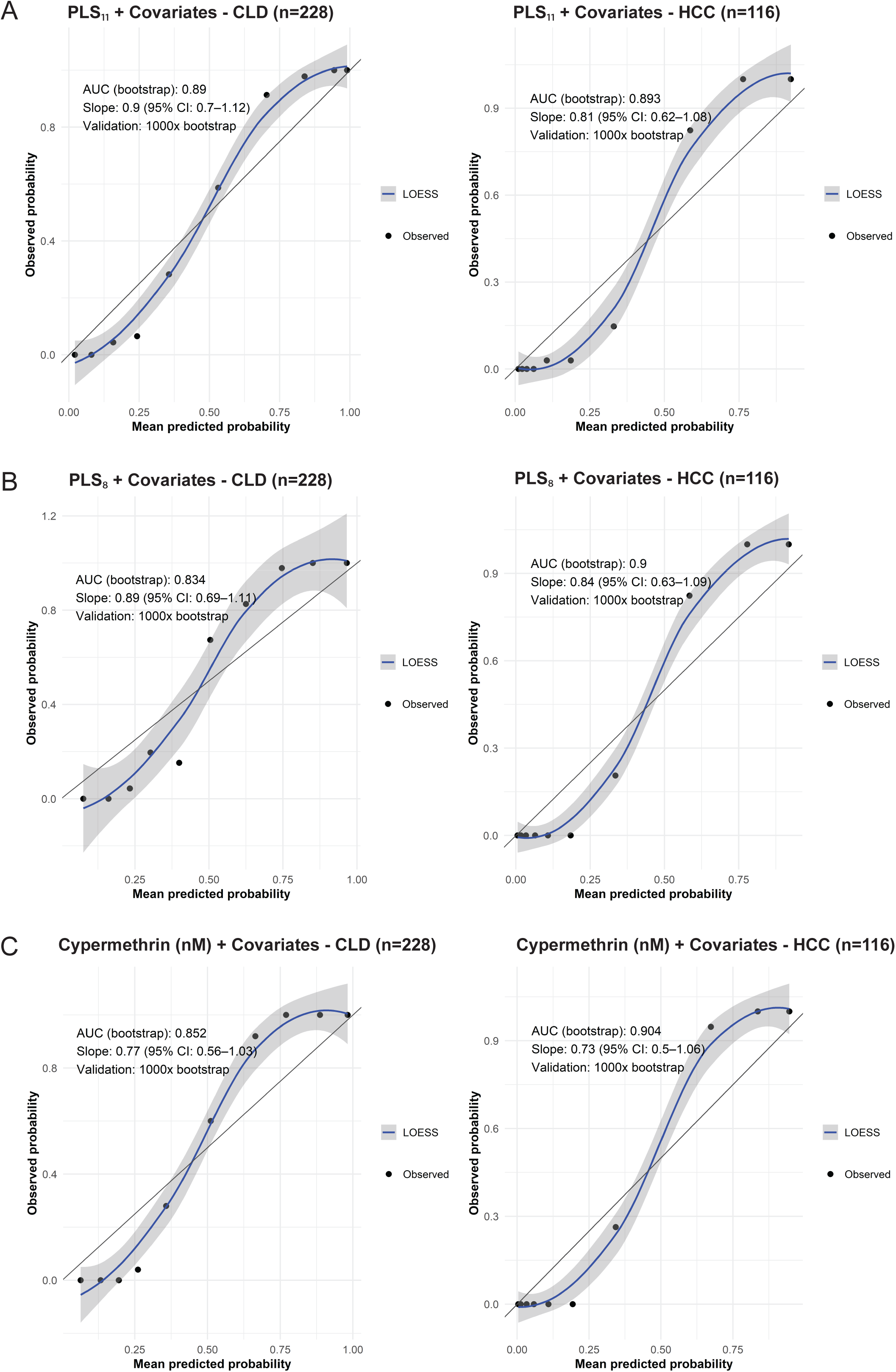
Calibration performance of PILCRS models with covariates. Calibration plots show observed versus predicted probabilities from logistic models incorporating PLS₁₁ (A), PLS₈ (B), or cypermethrin (nM) (C) plus covariates for CLD (left panels) and HCC (right panels), compared with HBC. Curves represent LOESS smoothing of observed probabilities; shaded areas show 95% CI. The black diagonal line indicates perfect calibration. AUC (bootstrap), slope (95% CI), and 1,000× bootstrap validation results are shown within each panel.

PILCRS₁₁ achieved the strongest performance. For CLD, it yielded an optimism-corrected AUC of 0ꞏ890 and calibration slope 0ꞏ900 (95% CI 0ꞏ705–1ꞏ118). For HCC, PILCRS₁₁ (AUC 0ꞏ893; slope 0ꞏ814) and PLS₈ (AUC 0ꞏ900; slope 0ꞏ840) both showed excellent discrimination and well-aligned calibration. Cypermethrin-based models retained predictive value (AUCs ≥0ꞏ85; slopes ≥0ꞏ70), though lower CI bounds for slope occasionally exceeded 1ꞏ0, indicating acceptable to good performance (appendix table S2, appendix p 4).

SHAP analysis decomposed predictor contributions across classifiers. In PLS₁₁ models, PLS₁₁ contributed most, followed by age, alcohol use, occupation, and sex. In PLS₈ models, PLS₈ and age were dominant, with alcohol use and occupation also important. In cypermethrin models, cypermethrin was primary, followed by alcohol use, age, occupation, and sex (appendix figure S5, appendix p 9). Contributions were directionally coherent and biologically plausible, supporting the framework’s relevance.

## Discussion

Leveraging biospecimens from the TIGER-LC hospital-based case–control study in Thailand— where pesticide exposure is extensive, monitoring infrastructure is still emerging, and HCC incidence is increasing—we found that urinary pesticide burden, particularly from PLS₁₁, PLS₈, and cypermethrin, was consistently higher in CLD and HCC cases than in hospital-based controls. Cypermethrin dominated the exposure distribution and, together with glyphosate-derived AMPA and PPA, emerged as a strong independent predictor across models.^9,14^

The magnitude of risk was substantial. Individuals with elevated PLS₁₁ exposure had nearly three-fold higher odds of CLD and five-fold higher odds of HCC compared with hospital-controls, while cypermethrin exposure was associated with up to a three-fold increase.^3^ Predicted probabilities also increased across exposure strata, with CLD probability approaching 70% in the highest quartile compared with below 50% in the lowest, and HCC probability roughly doubling. Predictive models, particularly PILCRS₁₁, achieved excellent discrimination and calibration,^27,28^ while gradient boosting further improved performance.^27^ SHAP analyses confirmed interpretability, with pesticides, age, and alcohol use contributing most strongly.^6^

These associations are biologically coherent and supported by mechanistic evidence. Cypermethrin induces mitochondrial dysfunction and NF-κB–mediated inflammation,^8,10,13^ while glyphosate analytes disrupt redox balance and DNA repair.^2,9,11^ Other pesticides—including pendimethalin, oxadiazon, metsulfuron-methyl, butachlor, 2,4-D, flocoumafen, and bromadiolone—exert overlapping hepatotoxic effects through oxidative stress, lipid peroxidation, and coagulation disruption.^6,7,15–17,19^ Supra-additive toxicities further justify integrative indices such as PLS₁₁ for capturing cumulative burden.^2,24–26^

Thailand, one of Southeast Asia’s largest pesticide consumers, lacks national biomonitoring and does not incorporate internal exposure into prevention policy.^20,22^ In this study, urinary concentrations of cypermethrin and glyphosate analytes frequently exceeded thresholds linked to hepatic injury.^9^ If externally validated, these findings have broad implications for LMICs with similar agrochemical practices and limited regulatory infrastructure,^21^ underscoring pesticide burden as a modifiable planetary health and environmental justice risk factor, disproportionately affecting rural and agricultural workers.^1,4^ Our exposure-informed MLRP framework addresses surveillance gaps by combining biospecimen-based exposure assessment with interpretable modelling.^27^ Direct quantification minimised recall bias, while bootstrap resampling strengthened internal validity.^28^

Limitations include a hospital-based case–control design that may introduce selection bias and restrict generalisability; single-spot urine exposure assessment with potential temporal variability, urine dilution from variable hydration, left-censoring at the limit of detection, and batch effects; an expanded yet incomplete pesticide panel with some non-specific metabolites; residual confounding from hepatitis B and C, alcohol, aflatoxin, diet, and metabolic risk factors; modest HCC and CLD sample sizes limiting subgroup analyses; and MLRP models validated internally only, lacking temporal and external validation to assess overfitting, transportability, calibration, and performance across population subgroups. Future studies should prioritise prospective cohorts with repeated urine sampling and temporal anchoring, incorporate untargeted exposomic screening, and integrate host-response data—including transcriptomics, immunophenotyping, and environmental DNA—to strengthen causal inference.^25,26^ With external validation, this modular MLRP framework could evolve into AI-enabled tools for population-level risk monitoring. Urinary pesticide profiling is minimally invasive and field-deployable,^2^ supporting integration into registries, occupational health programs, and WHO–FAO frameworks.^1,4^ By advancing a harmonized model grounded in biospecimen-derived data, reproducible MLRP, and environmental health informatics, this study delivers a scalable, policy-relevant solution aligned with the WHO Global Cancer Control Strategy, the IARC Cancer Prevention Roadmap, and SDG 3ꞏ9,^3,5,29,30^ contributing to an equity-driven planetary-health model that strengthens pesticide regulation, supports early prevention, and addresses environmental injustice in LMICs.

## Research in context Evidence before this study

Experimental studies show that pesticides such as glyphosate, 2,4-D, and cypermethrin cause hepatotoxicity through oxidative stress, mitochondrial dysfunction, and fibrogenic pathways. Epidemiological evidence remains limited and often relies on occupational or crop-type proxies prone to misclassification. Within TIGER-LC, we previously demonstrated that urinary glyphosate and its metabolites were elevated in patients with CLD and HCC compared with hospital-based controls, implicating pesticide exposure as a risk factor but restricted to single-compound analyses without cumulative or predictive modelling.

## Added value of this study

We extended urinary profiling to eight additional pesticides and derived a composite PLS to capture cumulative internal burden. This score was incorporated into two predictive models—a logistic regression–based PILCRS and an XGBoost classifier—across 593 TIGER-LC participants. Both demonstrated strong discrimination and calibration, with PILCRS achieving an optimism-corrected AUC of 0ꞏ91. CLD probability increased from 30% in the lowest quartile to more than 70% in the highest. Independent associations for cypermethrin and glyphosate metabolites were retained after adjustment for demographics and lifestyle. Sensitivity analyses confirmed robustness across subgroups, and calibration plots indicated close agreement between predicted and observed risks.

## Implications of all the available evidence

This study reframes pesticide burden as a modifiable determinant of liver disease, shifting from single-compound toxicology to cumulative, predictive stratification. The MLRP framework enables scalable surveillance and early prevention, adaptable to AI-enabled public health tools. Findings align with WHO–FAO priorities and support SDG 3ꞏ9, underscoring the urgency of regulation, biomonitoring, and protection for vulnerable populations in LMICs.

## Contributors

DPP and CAL conceptualized the study. DPP led the study design, data curation, and formal analysis. BP, SR, PN, NL, AC, CP, VL, AP, WS, TU, TS and KP contributed to data collection, validation, and interpretation. MH, MK, ALP, FJG, and AB contributed to manuscript review and editing. CM, XWW and MR provided supervision and scientific input. XW and CCH provided overall supervision, secured funding, and led project administration. DPP wrote the first draft of the manuscript. DPP, CAL, and XWW reviewed and interpreted the results. All authors reviewed and approved the final manuscript. DPP, CAL, and CCH were responsible for the decision to submit the manuscript for publication.

## Data sharing

The dataset used in this study will be made available to qualified researchers upon reasonable requests to the corresponding author, subject to institutional data use agreements and ethical approvals.

## Data Availability

All data produced in the present study are available from the corresponding author upon reasonable request.

## Acknowledgments

This research was supported [in part] by the Intramural Research Program of the National Institutes of Health (NIH). Funding was provided in part by the Intramural Research Program of the Center for Cancer Research, National Cancer Institute, US National Institutes of Health (ZIA BC 011492). The contributions of the NIH author(s) were made as part of their official duties as NIH federal employees, are in compliance with agency policy requirements, and are considered Works of the United States Government. However, the findings and conclusions presented in this paper are those of the author(s) and do not necessarily reflect the views of the NIH or the U.S. Department of Health and Human Services. Work in Thailand, including patient recruitment, data collection, and biospecimen banking, was supported by the Chulabhorn Research Institute, Bangkok, Thailand, and in part by Thailand Science Research and Innovation (TSRI) through the Chulabhorn Research Institute (grant 49890/4759784). We thank Vajarabhongsa Bhudhisawasdi, Chulabhorn Research Institute, Bangkok, and Khon Kaen University, Khon Kaen, Thailand; Chirayu U. Auewarakul, Chulabhorn Hospital, Bangkok, Thailand; and Suleeporn Sangrajrang, National Cancer Institute, Bangkok, Thailand, for their contributions to logistics, technical assistance, and data support.

## Abbreviations

2,4-D — 2,4-dichlorophenoxyacetic acid; AUC — area under the receiver operating characteristic curve; AI — artificial intelligence; AMPA — aminomethylphosphonic acid; BMI — body mass index; CI — confidence interval; CLD — chronic liver disease; DNA — deoxyribonucleic acid; FAO — Food and Agriculture Organization; FC — fold-change; GC–MS — gas chromatography– mass spectrometry; HBC — hospital-based controls; HBV — hepatitis B virus; HCC — hepatocellular carcinoma; HCV — hepatitis C virus; L1 — Lasso regularisation (penalises the absolute magnitude of coefficients, enabling feature selection); L2 — Ridge regularisation (penalises the squared magnitude of coefficients, retaining all features); LC–MS/MS — liquid chromatography–tandem mass spectrometry; LMICs — low- and middle-income countries; LOD limit of detection; LOQ — limit of quantification; MLRP — machine learning risk prediction; OR — odds ratio; PILCRS — Pesticide-Informed Liver Cancer Risk Score; PILCRS₈ — PLS₈-based score with clinical covariates; PILCRS₁₁ — PLS₁₁-based score with clinical covariates; PILCRSCYP — cypermethrin-based score with clinical covariates; PLS — Pesticide Load Score; PLS₈ — PLS based on eight urinary pesticide analytes; PLS₈ FC — fold-change in PLS₈, normalised to the control group median; PLS₁₁ — PLS based on eleven urinary analytes, including glyphosate and its metabolites (AMPA, PPA); PLS₁₁ FC — fold-change in PLS₁₁, normalised to the control group median; PPA — phosphoric acid; ROC — receiver operating characteristic; SD standard deviation; SDG — Sustainable Development Goal; SHAP — Shapley Additive Explanations; STROBE — Strengthening the Reporting of Observational Studies in Epidemiology; TIGER-LC — Thailand Initiative in Genomics and Expression Research for Liver Cancer; TRIPOD — Transparent Reporting of a Multivariable Prediction Model for Individual Prognosis or Diagnosis; TQ-S — triple quadrupole–sensitive; UPLC — ultra-performance liquid chromatography; WHO — World Health Organization; XGBoost — Extreme Gradient Boosting.

