## Appendix Supporting Information for "Urinary pesticide profiles and liver disease risk in Thailand: a machine-learning risk-prediction model"

**Targeted LC–MS/MS Assay for Cross-Sectional Quantification of Urinary Pesticides: Sample Preparation, Instrument Parameters, and Analytical Performance**

Urine samples were processed using a reversed-phase extraction protocol optimised for small-molecule pesticide detection. Briefly, 60 µL of urine was diluted 1:5 with extraction buffer (acetonitrile:water:methanol, 65:30:5, v/v) containing chlorpropamide (2 µM) as an internal standard. After centrifugation at 15 000 × g for 15 min, 250 µL of supernatant was transferred to a 96-well plate and diluted 1:2 with 50% methanol.

Target compounds were quantified using LC–MS/MS on a XEVO-TQSmicro triple quadrupole mass spectrometer (Waters Corporation, USA) with electrospray ionisation in both positive and negative modes. Chromatographic separation was performed on an ACQUITY UPLC BEH C18 column (1.7 µm, 50 × 2.1 mm) maintained at 40 °C, with a 5 µL injection volume. Source parameters included a capillary voltage of 2.39 kV, source temperature of 150 °C, desolvation temperature of 500 °C, and desolvation gas flow of 1000 L/h. Data acquisition was performed in multiple reaction monitoring (MRM) mode with transitions optimised using IntelliStart.

The following precursor–product ion transitions (m/z), cone voltages (V), and collision energies (eV) were used: pendimethalin, 282.11 → 212.04 (16, 8); oxadiazon, 345.04 → 219.97 (42, 18); metsulfuron-methyl, 141.10 → 42.68 (72, 18); butachlor, 312.13 → 238.05 (18, 10); 2,4-dichlorophenoxyacetic acid (2,4-D), 218.95 → 160.89 (28, 12); cypermethrin, 415.98 → 190.88 (48, 12); flocoumafen, 543.20 → 355.17 (56, 22); and bromadiolone, 525.11 → 250.04 (48, 36). Chlorpropamide (277.07 → 110.96; 28, 30) was used as the internal standard. Quantification was performed using external calibration curves generated from authentic standards in 50% methanol.

Analytical validation showed signal enhancement as the predominant matrix effect across compounds. Limits of detection ranged from 0.16 to 467 nM, and limits of quantification were ≤0.5 nM for six of eight pesticides. Extraction efficiency exceeded 200% across concentration levels, with reproducible recovery and minimal ion suppression. Detection frequencies in the study population ranged from 3.7% (bromadiolone) to 98.6% (cypermethrin), supporting high assay sensitivity under field conditions (Table S1).

**Table S1: Targeted LC–MS/MS assay for cross-sectional quantification of urinary pesticides: compound use profiles, mass spectrometry parameters, and analytical performance**

| ISO common name | Use type | Target pest or weed | Matrix effect | Suppression / Enhancement ratio | LOD (nM) | LOQ (nM) | Extraction efficiency (high, %) | Extraction efficiency (medium, %) | Extraction efficiency (low, %) | Detection frequency (*n* samples) |
|---|---|---|---|---|---|---|---|---|---|---|
| Pendimethalin | Herbicide | Grasses, broadleaf weeds (microtubule inhibitor) | Enhancement | 1·47 | 0·16 | 0·5 | 1810·03 | 2009·83 | 270·04 | 55 |
| Oxadiazon | Herbicide | Grasses, some broadleaves (PPO inhibitor) | Enhancement | 1·416 | 0·5 | 0·5 | 412·60 | 435·90 | 534·82 | 66 |
| Metsulfuron-methyl | Herbicide | Broadleaf weeds (ALS inhibitor) | Enhancement | 2·14 | 467 | 0·5 | 233·73 | 328·17 | 380·89 | 649 |
| Butachlor | Herbicide | Annual grasses in rice (pre-emergent) | Enhancement | 1·416 | 0·5 | 13·67 | 341·29 | 340·95 | 395·07 | 474 |
| 2,4-D | Herbicide | Broadleaf weeds (auxin mimic) | Enhancement | 1·47 | 467 | 467 | 252·13 | 240·29 | 231·23 | 24 |
| Cypermethrin | Insecticide | Broad-spectrum (pyrethroid) | Enhancement | 24 | 370·3 | 370·3 | 354·09 | 315·57 | 326·50 | 1085 |
| Flocoumafen | Rodenticide | Rodents (anticoagulant) | Enhancement | 4·48 | 0·167 | 4·67 | 747·32 | 923·91 | 938·46 | 601 |
| Bromadiolone | Rodenticide | Rodents (rats, mice; anticoagulant) | Enhancement | 1·438 | 41 | 41 | 280·98 | 268·80 | 209·18 | 22 |

ISO = International Organization for Standardization. Urine matrix effects predominantly resulted in signal enhancement, indicating that ion suppression was not a major contributor to low detection. All pre-spiked pesticides were efficiently recovered, with extraction inefficiency as the main limiting factor. LOD = lowest concentration with signal-to-noise ratio (SNR) ≥3 (RMS); LOQ = lowest concentration with SNR ≥10 (RMS).

### Table S2: Performance characteristics and calibration of internal exposure-based liver disease risk models

| Model | Outcome | Corrected AUC | Calibration slope | 95% CI (slope) | Calibration interpretation | Thresholds for performance classification |
|---|---|---|---|---|---|---|
| **$PILCRS_{11}$** | CLD | 0·890 | 0·900 | 0·705–1·118 | Excellent | AUC ≥ 0·85 and Slope 0·85–1·15 |
| **$PILCRS_{11}$** | HCC | 0·893 | 0·814 | 0·617–1·079 | Good | AUC ≥ 0·85 and Slope 0·75–1·15 |
| **$PILCRS_{8}$** | CLD | 0·834 | 0·889 | 0·689–1·110 | Good | AUC ≥ 0·80 and Slope 0·85–1·15 |
| **$PILCRS_{8}$** | HCC | 0·900 | 0·840 | 0·627–1·095 | Excellent | AUC ≥ 0·85 and Slope 0·80–1·15 |
| **$PILCRS_{CYP}$** | CLD | 0·852 | 0·772 | 0·563–1·031 | Good | AUC ≥ 0·85 and Slope 0·75–1·15 |
| **$PILCRS_{CYP}$** | HCC | 0·904 | 0·734 | 0·504–1·060 | Acceptable | AUC ≥ 0·85 and Slope ≥ 0·70 with CI crossing 1·0 |

$PILCRS_{11}$ and $PILCRS_{8}$ denote pesticide-informed liver cancer risk scores derived from logistic regression models incorporating $PLS_{11}$ or $PLS_{8}$ scores and clinical covariates, respectively. $PILCRS_{CYP}$ refers to a cypermethrin-based model. AUC indicates area under the receiver operating characteristic curve; values closer to 1·0 indicate superior discrimination. Calibration slope values near 1·0 indicate good model calibration. 95% CI = 95% confidence interval. Internal validation performed using 1000× bootstrap resamples.

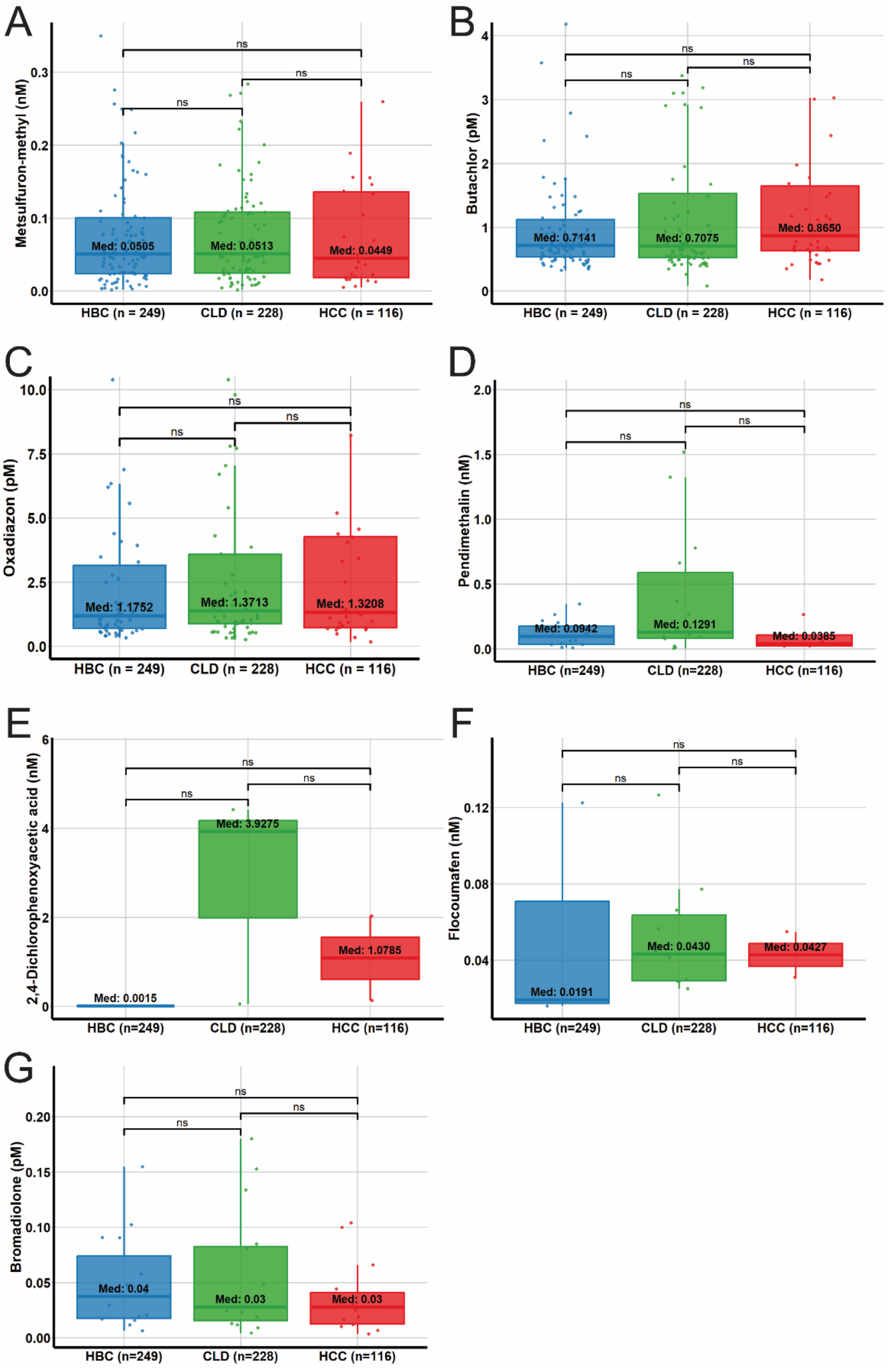


**Figure S1:** Distribution of urinary pesticide concentrations across disease groups. (A–G) Boxplots show creatinine-adjusted concentrations of seven pesticides—(A) metsulfuron-methyl, (B) butachlor, (C) oxadiazon, (D) pendimethalin, (E) 2,4-D, (F) flocoumafen, and (G) bromadiolone—across HBC ($n$ = 249), CLD ($n$ = 228), and HCC ($n$ = 116). Medians are shown within boxes; horizontal bars denote pairwise comparisons. *ns* = non-significant by Wilcoxon rank-sum test. HBC = hospital-based controls.

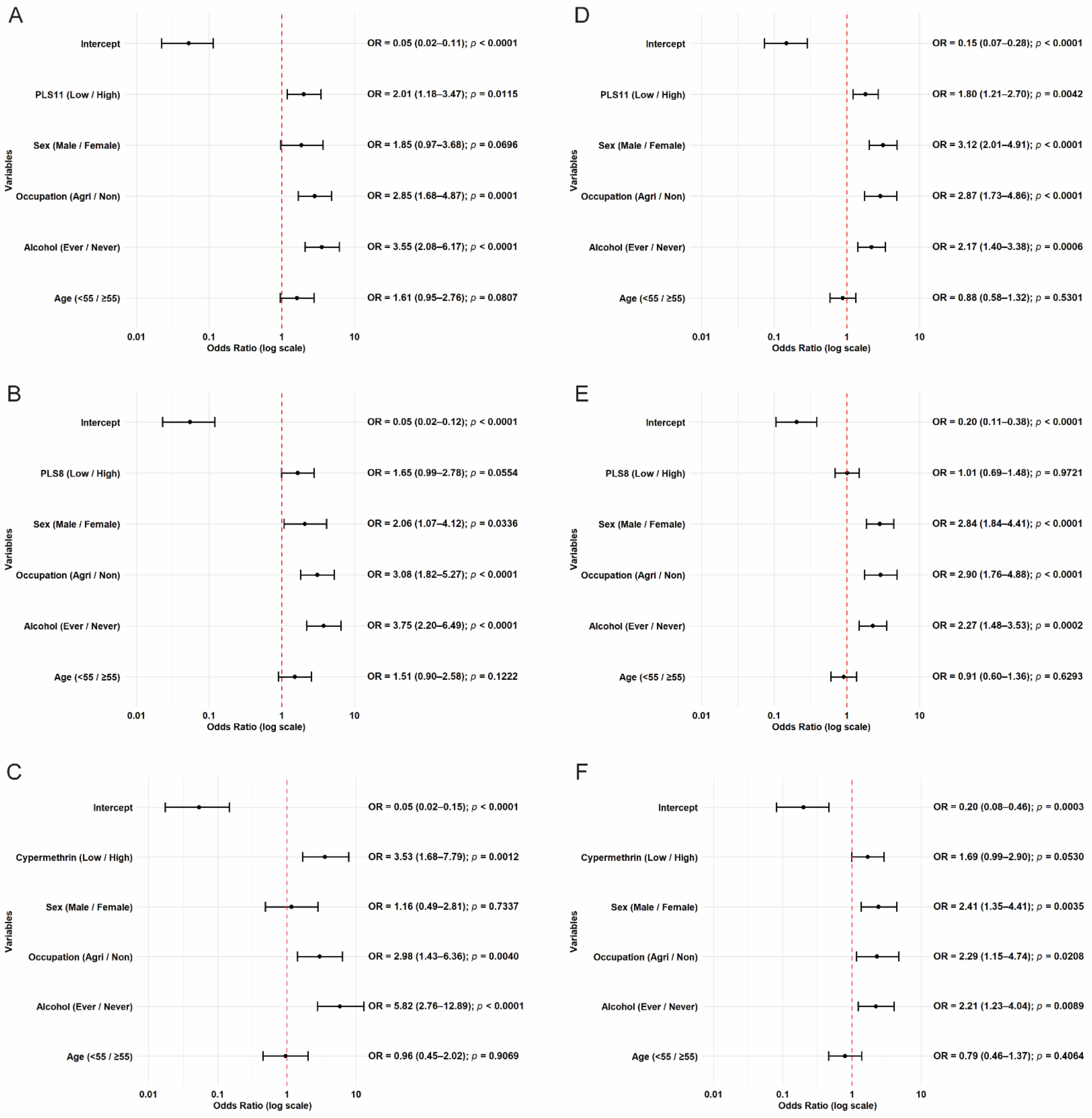


**Figure S2: Multivariable-adjusted associations of pesticide exposure and PLS scores with liver disease.** (A–C) Forest plots show adjusted odds ratios (ORs, log scale) for CLD versus HBC based on exposure to $PLS_{11}$ (A), $PLS_{8}$ (B), and cypermethrin (C), controlling for sex, occupation, alcohol use, and age. (D–F) Corresponding models for HCC versus HBC. Point estimates and 95% confidence intervals are shown; p values are derived from logistic regression models. HBC = hospital-based controls.

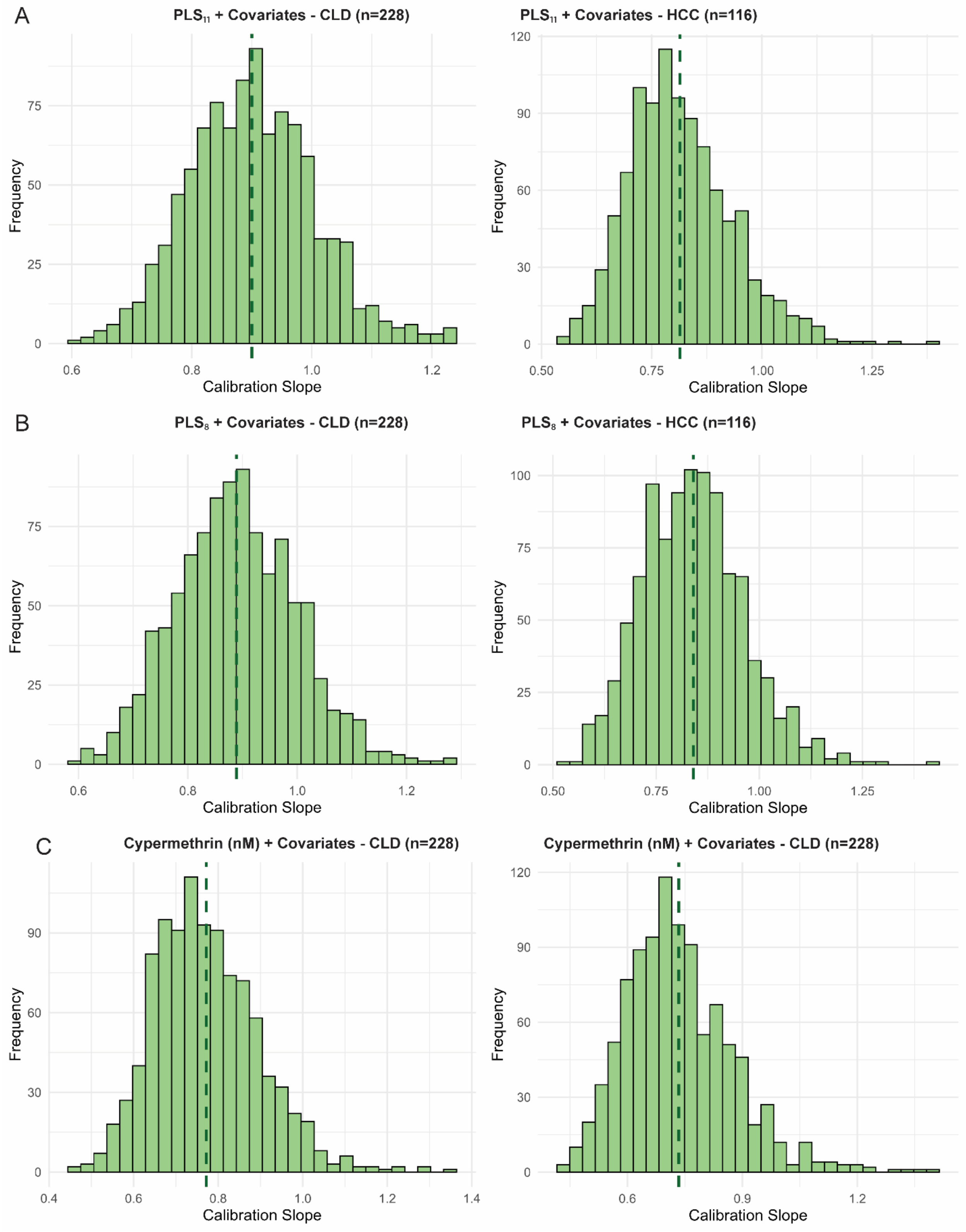


**Figure S3: Internal calibration of predictive models for liver disease.** (A–C) Histograms show calibration slope distributions from 1000 bootstrap resamples for models predicting CLD (left) and HCC (right) using $PLS_{11}$ (A), $PLS_8$ (B), and urinary cypermethrin (nM) (C), adjusted for age, sex, occupation, and alcohol use. Dashed lines indicate the median slope; values near 1·0 indicate stronger calibration.

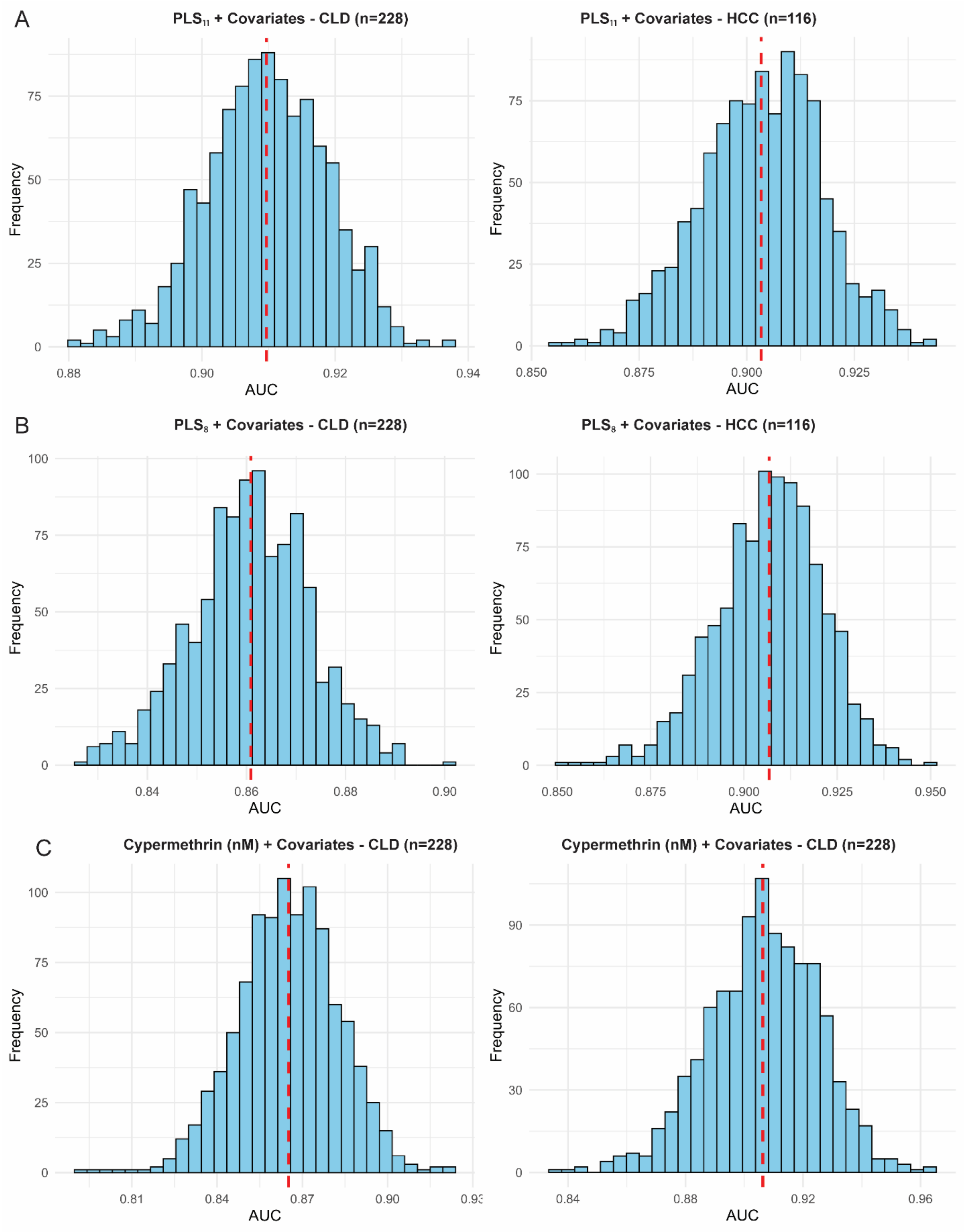


**Figure S4: Internal validation of model discrimination.** (A–C) Histograms show AUC distributions from 1000 bootstrap resamples for models predicting CLD (left) and HCC (right) using $PLS_{11}$ (A), $PLS_{8}$ (B), and urinary cypermethrin (nM) (C), adjusted for age, sex, occupation, and alcohol use. Red dashed lines indicate the median AUC; higher values indicate better discrimination between cases and controls.

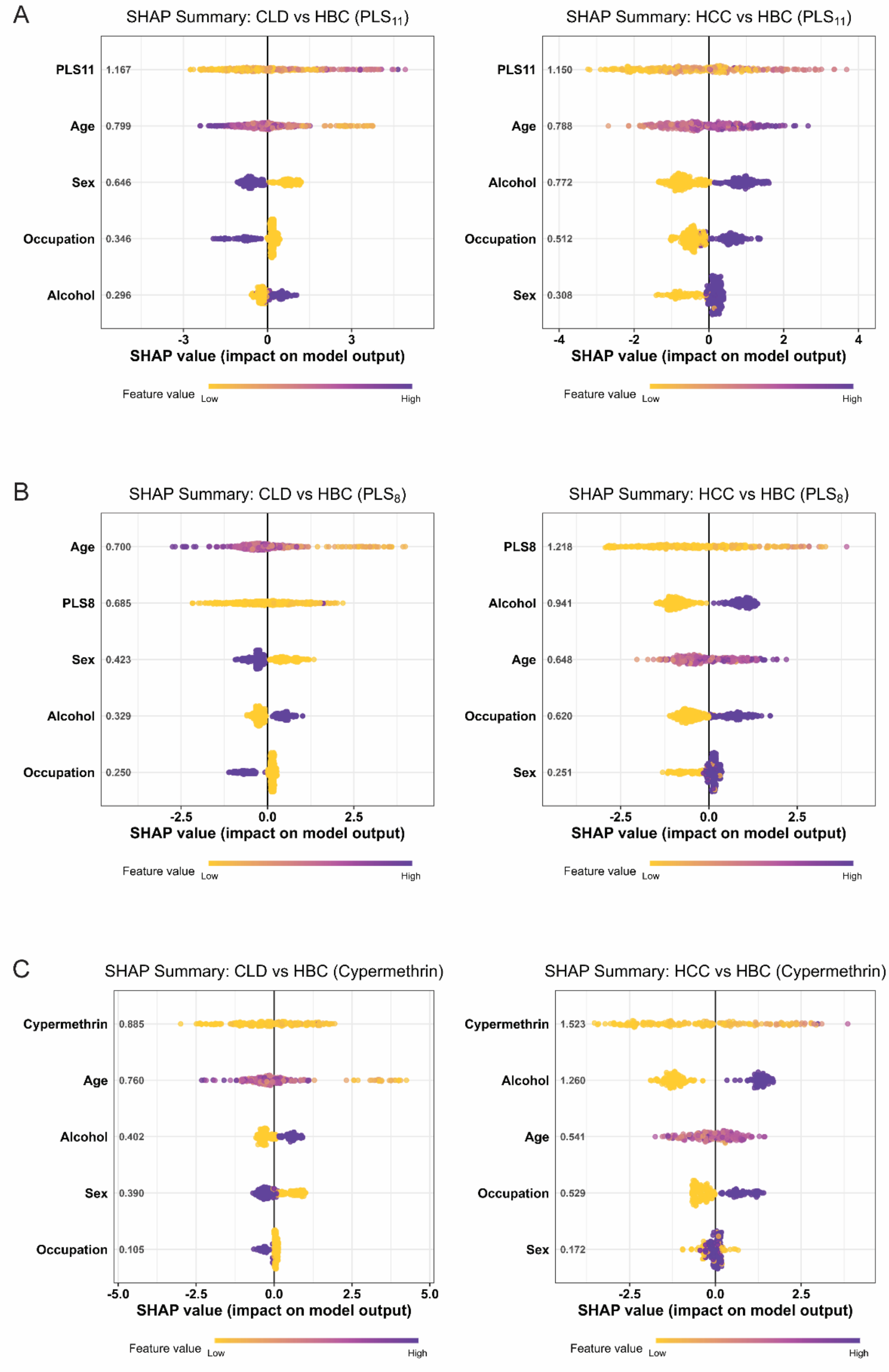


**Figure S5: SHAP summary plots for liver disease risk.** SHAP plots show feature contributions for CLD (left) and HCC (right) using $PLS_{11}$ (A), $PLS_8$ (B), and urinary cypermethrin (nM) (C), adjusted for age, sex, occupation, and alcohol use. Features are ranked by mean absolute SHAP value; colour gradients indicate scaled feature expression from low (yellow) to high (purple). Models were trained separately for CLD versus HBC and HCC versus HBC. HBC = hospital-based controls.
